# Prevalence and determinants of modern contraception use among persons with disabilities in low- and middle-income countries: a systematic review and meta-analysis

**DOI:** 10.1101/2024.09.13.24313669

**Authors:** Atika Rahman Chowdhury, Shimlin Jahan Khanam, Mohammad Zahidul Islam, Gulam Khandaker, Md Nuruzzaman Khan

## Abstract

**Background:** Persons with disabilities should require the same level of access to contraception as the general population. However, the extent of contraception use among this group is underexplored in low- and middle-income countries (LMICs).

**Objective:** This study aimed to determine the prevalence of modern contraception use among persons with disabilities in LMICs and identify the key determinants.

**Methods:** In June 2024, we conducted a systematic search across six databases to identify studies on disability and modern contraception in LMICs. The primary outcomes were the prevalence of modern contraception use and its determinants among persons with disabilities. Summary estimates were calculated using fixed or random-effects meta-analysis, depending on the level of heterogeneity.

**Results:** A total of 19 studies were identified, with 11 included in the meta-analysis. The pooled prevalence of modern contraception use among persons with disabilities was 31.4% (95% CI: 26.5, 36.2), with significant heterogeneity across respondent characteristics. Five factors were significantly associated with higher contraception use: age over 25 years, having some level of education, being in a higher wealth quintile, adequate knowledge of family planning, and being in a formal marital relationship.

**Conclusion:** This study reveals a significantly lower prevalence of modern contraception use among persons with disabilities in LMICs. Improving access to education, addressing social norms, and strengthening healthcare systems may contribute to increase contraception access and uptake among persons with disabilities in LMICs.

## Background

An estimated 1.3 billion people, representing 16% of the global population, are currently living with disabilities ^1^. Over 80% of them reside in low- and middle-income countries (LMICs), majority of those without access to rehabilitation, education and other essential health services ^2^. The majority of individuals with disabilities in LMICs rely on governmental and social support to meet their basic needs due to limited education and employment opportunities, as well as the persistent perception of them as burdens to society ^3-5^. In this context, ensuring their well-being is particularly challenging, as community efforts often focus solely on providing basic necessities like food and clothing ^6^. Access to healthcare services is also restricted by factors such as lack of disability-friendly healthcare facilities, inadequate transportation options, and the costs associated with accessing care ^2, 7^.

The challenges of accessing healthcare services for persons with disabilities are particularly pronounced in the area of sexual and reproductive health, including family planning and contraception ^7-9^. The underlying reasons are multifaceted, including community assumptions about their limited or non-existent sexual lives, taboos related to family planning and contraception that are prevalent even among the general population in low resource settings, and the neglect of government family planning providers in existing services ^10-13^. These issues persist despite evidence in the literature that individuals with disabilities lead sexual lives similar to those of non-disabled people ^14, 15^. Consequently, persons with disabilities experience higher rates of unintended and closely spaced pregnancies, which further increase their vulnerability to adverse pregnancy outcomes and poor health ^16-18^. Additionally, as people with disabilities often live in poverty, their children are more likely to remain trapped in the cycle of poverty, leading to issues such as school dropout and early marriage, perpetuating a cycle of disadvantage ^19-21^. These burdens directly challenge LMICs in achieving the Sustainable Development Goals (SDGs), particularly the goal of “leaving no one behind” and ensuring equitable access to sexual and reproductive health under SDG 3 ^22^.

Addressing the challenges of ensuring family planning and contraception for persons with disabilities requires international and national policies and programs that focus on providing these services at the household level. This effort necessitates accurate estimates of contraception use among persons with disabilities in LMICs through comprehensive research. However, this aspect is particularly challenging in LMICs, as existing research primarily focuses on contraception use and its determinants among the general population ^23^, often systematically excluding persons with disabilities due to community-level ignorance of this issue ^24^. Moreover, research in LMICs often relies on secondary data sources, such as the Demographic and Health Survey and the Multiple Indicator Cluster Survey, which do not specifically address persons with disabilities ^25^. Consequently, the limited evidence available is based on small surveys with less precise analysis and inadequate consideration of a broad range of confounders. As a result, there are no accurate estimates of contraception use among persons with disabilities in LMICs, comparable to those available for the general population. A systematic review and meta-analysis of existing studies can help address this gap. Therefore, we conducted this study to summarize the prevalence of modern contraception use among persons with disabilities in LMICs and to identify their major determinants.

## Methods

We conducted a systematic review and meta-analysis, following the Preferred Reporting Items for Systematic Reviews and Meta-Analyses (PRISMA) guidelines for observational studies. We included relevant studies on contraception use among persons with disabilities.

### Search Strategy

In June 2024, we conducted a thorough literature search in six databases: PubMed, Web of Science, Embase, Global Health, Medline, and Scopus (Supplementary table 01-06). We included studies published from January 2015 to June 2024, aligning with the establishment of the SDGs. Our search strategies included various keywords and medical subject headings (MeSH) related to the exposure, outcomes, and settings of interest. We also searched the websites of selected journals and reviewed reference lists of relevant articles.

### Inclusion Criteria

To be eligible for inclusion, articles had to meet the following criteria: (i) peer-reviewed journal articles, (ii) written or published in English, (iii) published from 2015 onwards, (iv) reported information on modern contraception use among persons with disability, (v) conducted in any LMICs, and (vi) presented original research using quantitative, qualitative, or mixed-methods.

### Exclusion Criteria

We excluded studies involving: (i) reproductive-aged women without disabilities, (ii) reproductive-aged women with high-risk characteristics like HIV/AIDS or cancer, (iii) studies conducted outside LMICs, (v) articles published before 2015, and (vi) studies published in the form of conference presentations, student theses, editorials, letters to the editor, commentaries, symposium proceedings, or those that were not peer-reviewed.

### Study Selection

Two authors (XX and XX) independently reviewed all articles, starting with title and abstract screening. Articles selected at this stage underwent full-text review by the same authors (XX and XX). Any disagreements were resolved through discussion, involving the lead author (XX) if necessary. We used online platforms like COVIDENCE, EndNote 21, and face-to-face meetings for efficient collaboration during the review process.

### Data Extraction

Before tabulating the final data, we designed a data extraction template, trialed and modified. We extracted relevant data, including authors’ names, study design, sample size, study setting, related exposure groups, and family planning or contraceptive uptake information. Additionally, we extracted reported effect sizes (odds ratios, ORs) and the underlying data used to calculate them. We also noted whether the ORs were adjusted or unadjusted for possible confounders. If there were any disagreements between the data extractors, we resolved them through discussion, involving the lead author (XX) if necessary.

### Quality assessment

We used the modified Newcastle-Ottawa Scale (NOS) for quality assessment of the included studies. The items included in the scale varied depending on the study type (cross-sectional, case-control, cohort studies, or randomized control trials). Two authors (XX and XX) assessed the included articles and assigned 1 point for each item if the study met the relevant criteria. We used aggregated scores to measure the overall study quality, categorizing it as good (score: 8 to 9), moderate (score: 5 to 7), or low (score: <5) following the guidelines.

### Study variables

The study variables were modern contraception use among persons with disabilities and their determinants. We classified contraception according to the World Health Organization’s guidelines on modern contraception. They include: hormonal contraceptive methods (oral pills or implants, injectables, patches or vaginal rings), intrauterine devices (IUDs), male and female condom, male and female sterilization ^26^.

### Statistical analysis

We used the extracted odds ratios (ORs) as the basis for our calculations. If the ORs were not available in the paper, we calculated unadjusted ORs based on the underlying data. The summary estimate of contraception prevalence among persons with disabilities were determined through fixed-effects or random-effects models determined heterogeneity assessment using the *I*^2^ statistic with a p-value. Random-effects model was considered where reported heterogeneity was moderate (50-74%) or high (75-100%). We also conducted subgroup analysis and meta-regression to explore the source of heterogeneity once we identified moderate or higher heterogeneity. The pre-specified subgroups included study sample size, confounding adjustment, study design, study settings, and related exposure groups. To assess publication bias, we visually inspected the Funnel Plot for asymmetry and conducted Egger’s regression test. For all analyses, we used the statistical software STATA version 16.1. The manuscript is written as per STROBE checklist (supplementary file 1)

## Results

### Search results

We identified a total of 314 articles from six databases and an additional 95 articles through searches on relevant journal websites, Google, Google Scholar, and reference lists of selected articles. After removing 166 duplicates, 243 unique articles remained. Screening of titles and abstracts led to the exclusion of 134 studies. Full-text reviews were conducted on the remaining 39 articles, with 21 further excluded based on the full-text review (Supplementary Table 7). Ultimately, 18 studies were included in our analysis: 11 in the quantitative synthesis and 7 in the narrative synthesis. The included studies were of moderate to good quality (Supplementary Table 8-9).

### Study Characteristics

The total sample size analyzed across the 18 included studies was 313,983 persons with disability. The majority of the studies were conducted in Ethiopia (n=7), followed by Uganda (n=2), Nigeria (n=2), Kenya (n=1), India (n=1), Ghana (n=1), Senegal (n=1), Pakistan (n=1), Bangladesh (n=1), and Nepal (n=1) (Supplementary Table 07). Of these studies, 16 employed a cross-sectional design, and almost two-thirds (n=11) reported community-based data.

### Prevalence of Modern Contraception Among Persons with Disabilities

Out of the 18 studies included in the review, 11 reported the prevalence of modern contraception use, with a combined sample size of 58,895 persons with disability. Modern contraception use among persons with disabilities in LMICs varied across studies, ranging from 20% to 49% ^27, 28^. The summary estimate, calculated using a random-effects meta-analysis, yielded an average prevalence of 31.4% (Figure 2).

**Figure 1.**
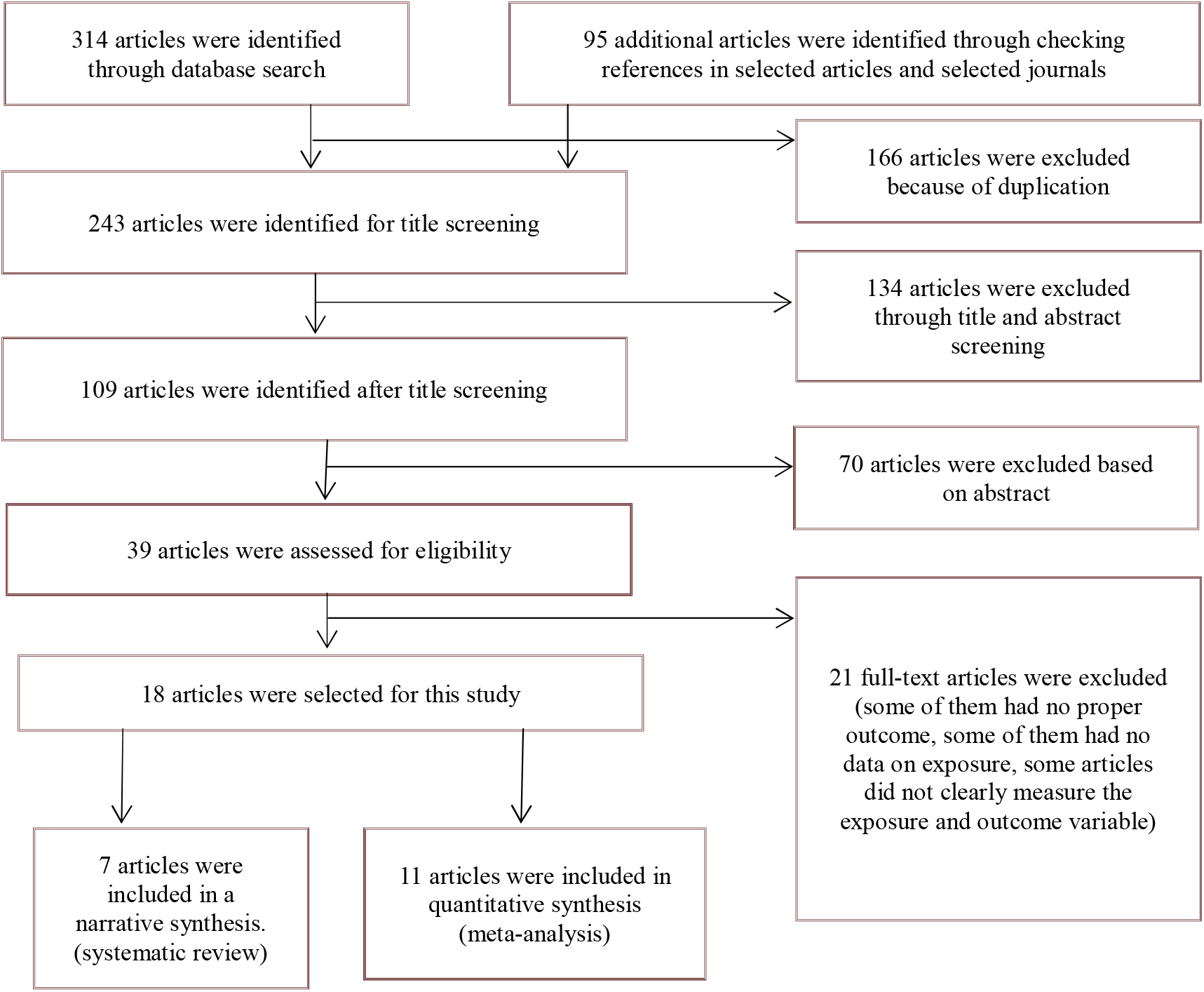
Schematic presentation of the studies included in the systematic review

**Figure 2.**
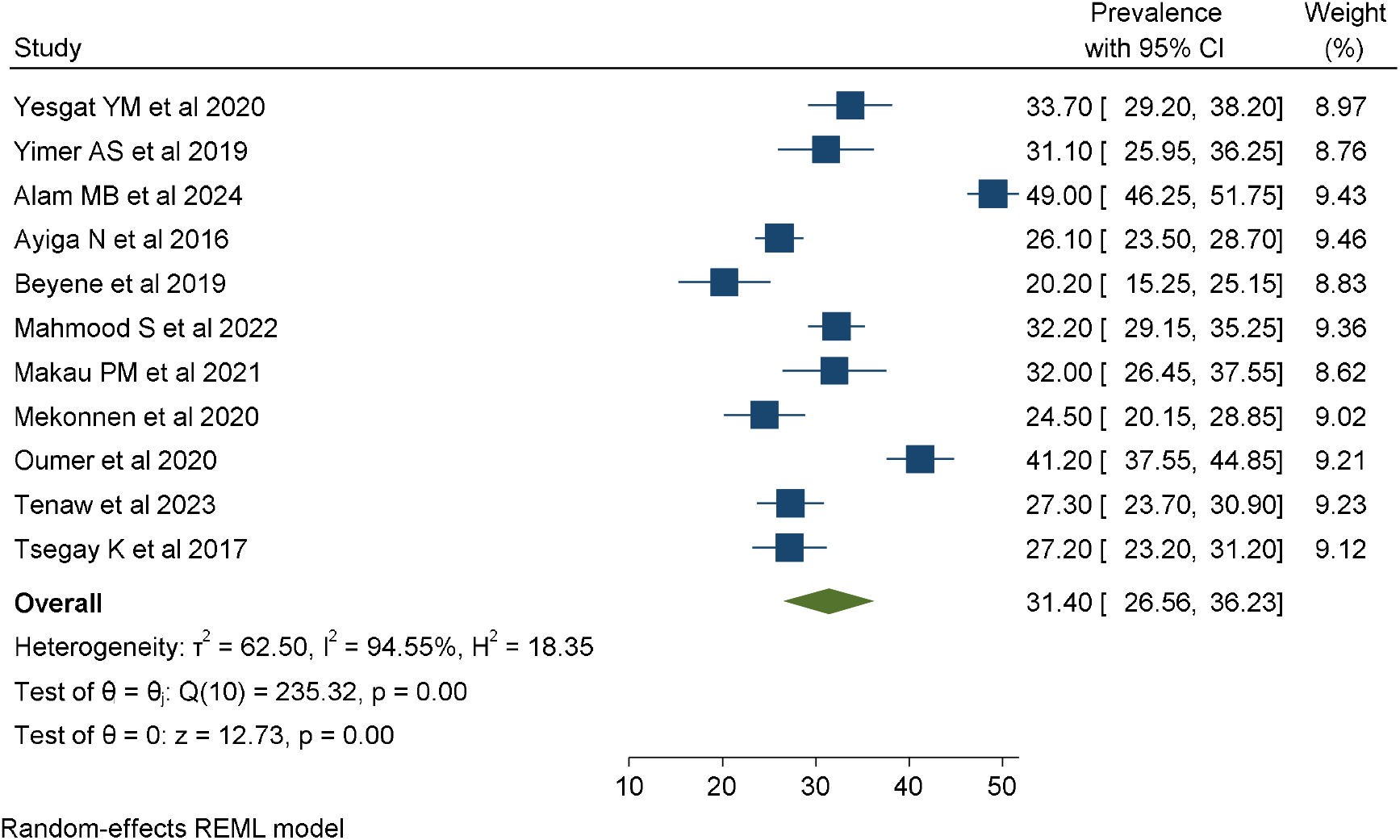
Summary estimation of prevalence of modern contraception use among persons with disabilities in low- and middle-income countries, January 2015 to June 2024.

**Figure 3.**
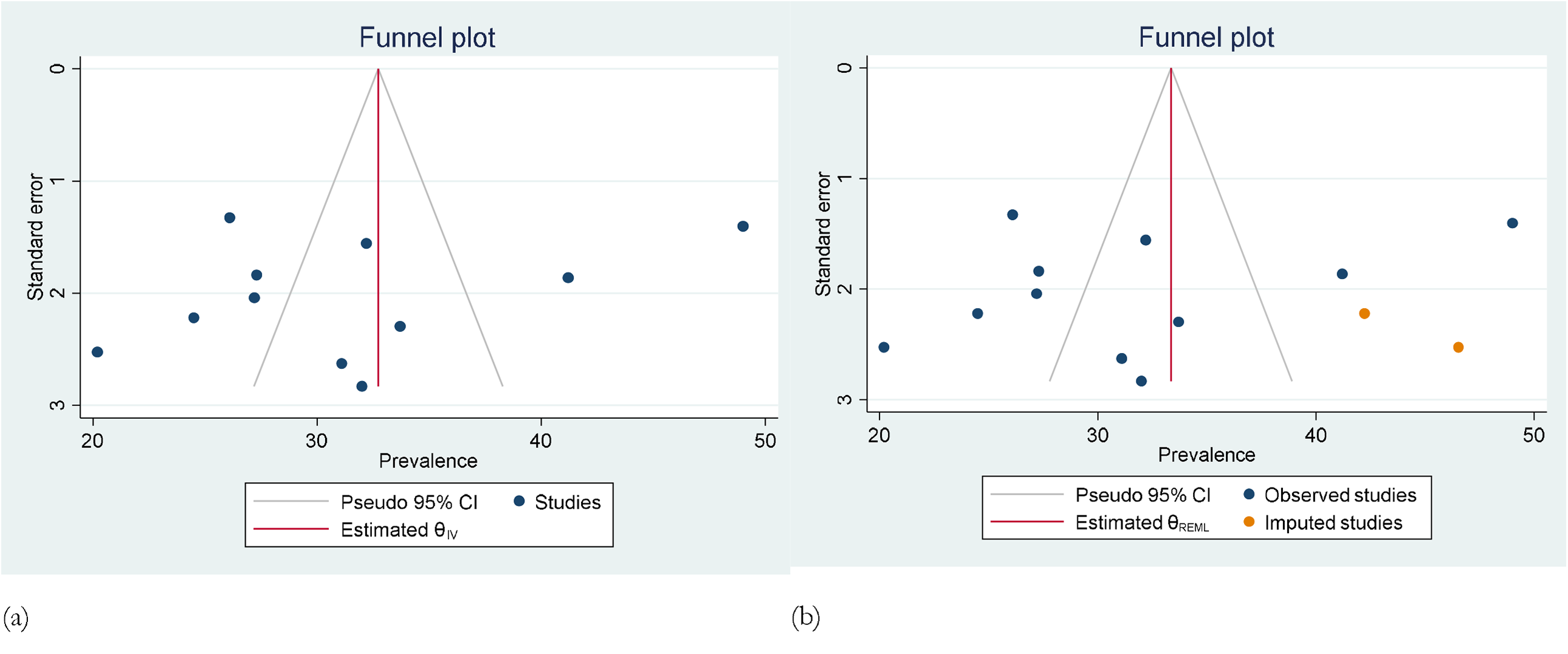
Funnel plots to access publication bias (a) and addressing publication bias through trim and fill estimate.

### Publication bias and estimate after adjustment

We found evidence of publication bias in the overall prevalence estimate. Adjusting for this bias using the trim and fill method identified two missing studies. However, including these studies in the analysis did not result in any significant change in the estimated prevalence of contraception.

### Subgroup Analysis and Meta-Regression

We observed significant heterogeneity in the estimated prevalence of modern contraception. To identify the sources of this heterogeneity, we conducted subgroup analyses based on sample size, study setting, study design, and country. The overall prevalence of modern contraception use was estimated at 40.61% (95% CI: 24.15-57.08) in studies with sample sizes of 5,352 or more and 35.77% (95% CI: 22.33-49.20) in national-level studies (Table 1). The highest prevalence of modern contraception use was reported among persons with disabilities in Bangladesh, at 49.00% (95% CI: 46.25-51.75). The corresponding forest plots are presented in the supplementary figure 1-4.

**Table 1:**
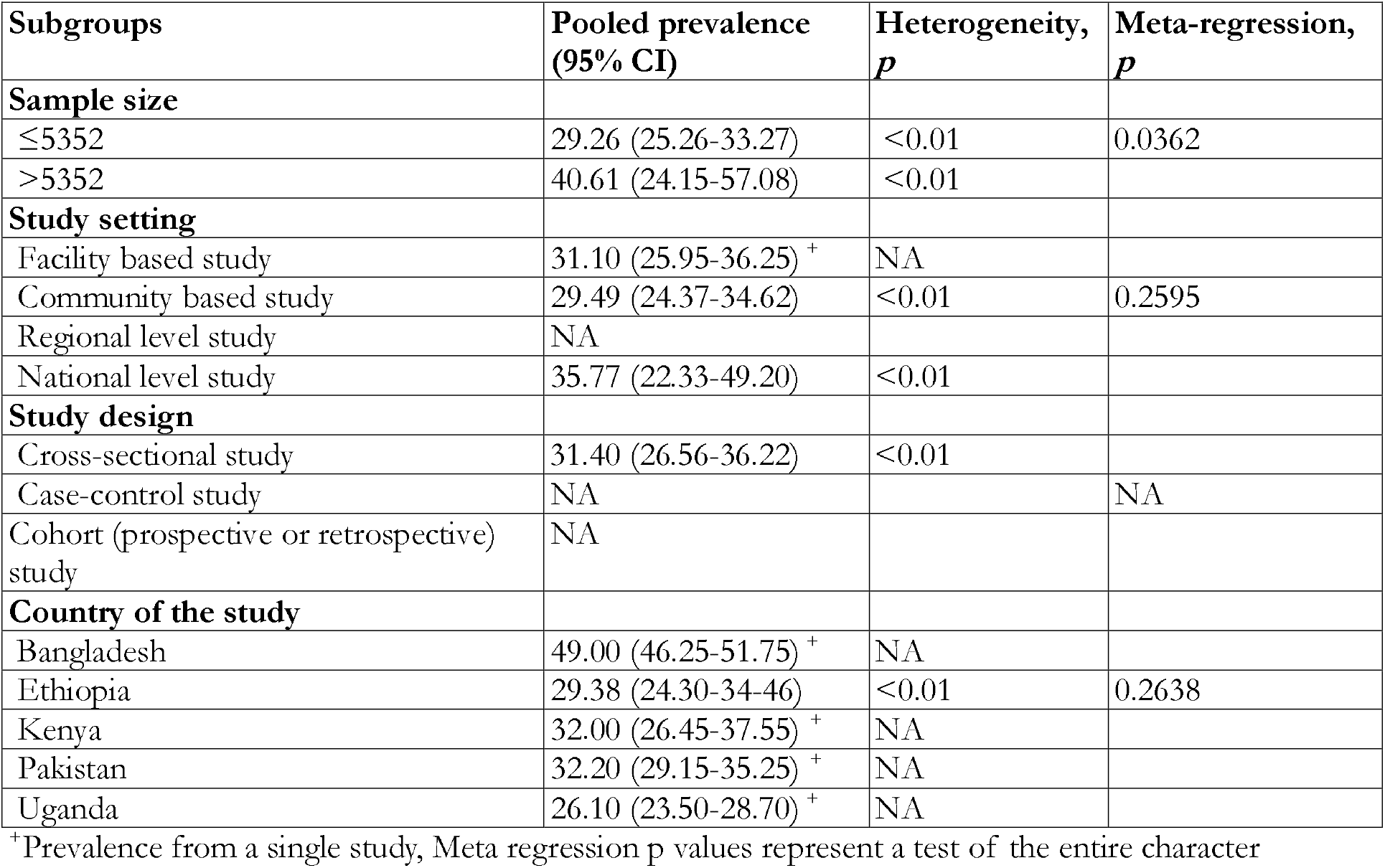
Pooled prevalence of contraceptive use among persons with disability in low- and middle-income countries across several selected characteristics, January 2015 to June 2024.

| Subgroups | Pooled prevalence<br>(95% CI) | Heterogeneity,<br>*p* | Meta-regression,<br>*p* |
| --- | --- | --- | --- |
| **Sample size** |  |  |  |
| ≤5352 | 29.26 (25.26-33.27) | <0.01 | 0.0362 |
| >5352 | 40.61 (24.15-57.08) | <0.01 |  |
| **Study setting** |  |  |  |
| Facility based study | 31.10 (25.95-36.25) <sup>+</sup> | NA |  |
| Community based study | 29.49 (24.37-34.62) | <0.01 | 0.2595 |
| Regional level study | NA |  |  |
| National level study | 35.77 (22.33-49.20) | <0.01 |  |
| **Study design** |  |  |  |
| Cross-sectional study | 31.40 (26.56-36.22) | <0.01 |  |
| Case-control study | NA |  | NA |
| Cohort (prospective or retrospective) study | NA |  |  |
| **Country of the study** |  |  |  |
| Bangladesh | 49.00 (46.25-51.75) <sup>+</sup> | NA |  |
| Ethiopia | 29.38 (24.30-34.46) | <0.01 | 0.2638 |
| Kenya | 32.00 (26.45-37.55) <sup>+</sup> | NA |  |
| Pakistan | 32.20 (29.15-35.25) <sup>+</sup> | NA |  |
| Uganda | 26.10 (23.50-28.70) <sup>+</sup> | NA |  |

<sup>+</sup>Prevalence from a single study, Meta regression p values represent a test of the entire character

### Factors associated with modern contraception use among persons with disabilities in low- and middle-income countries

We found that the 11 included studies identified a total of 19 variables significantly associated with modern contraception use among persons with disabilities. These variables were: age (n=6) ^9, 13, 27, 29-31^, education (n=5) ^9, 13, 27, 29, 30^, occupation (n=4)^9, 13, 27, 29^, wealth index (n=5) ^9, 13, 27, 29, 30^, attitude towards contraception (n=5) ^9, 13, 31-33^, knowledge about contraception (n=4) ^9, 13, 32, 34^, marital status (n=6) ^9, 13, 27, 30, 32, 34^, proximity to healthcare facilities (n=2) ^9, 28^, residence (n=1) ^9^, discussion about contraception with a partner or family members (n=1) ^31^, partner’s educational status (n=1) ^29^, partner’s economic status (n=1) ^29^, availability of transportation (n=1) ^9^, parity (n=1) ^13^, antenatal care visits (n=1) ^30^, availability of friendly sexual and reproductive healthcare services (n=1) ^13^, living children (n=1) ^30^, difficulty in accessing health services (n=1) ^30^, and confidentiality/privacy at health facilities (n=1) ^31^.

Of these, eight variables were reported as significant determinants of modern contraception use in at least two studies. Their summary effect sizes are provided in Table 2. In summary estimate of the significant variables, five variables were found significant: age over 25 years (OR, 2.26, 95% CI: 1.27-4.02), having education (OR, 1.94, 95% CI: 0.91-1.83), being in a wealth quintile other than poor (OR, 1.95, 95% CI: 1.18-3.23), possessing adequate knowledge about contraception (OR, 3.23, 95% CI: 1.57-6.62), and being in a formal marital relationship (OR, 3.32, 95% CI: 1.59-6.90). The corresponding forest plots are presented in the supplementary figure 5-12.

**Table 2:** Major factors associated with utilization of modern contraception among persons with disabilities in low- and middle-income countries, January 2015 to June 2024.

| **Determinants** | **Reference category** | **No. of studies** | **Sample size** | **OR (95%CI)** | **P-value** | **I<sup>2</sup> (%)** | **Heterogeneity test (P-value)** |
| --- | --- | --- | --- | --- | --- | --- | --- |
| Age | ≤ 25 | 6 | 3649 | 2.26 (1.27-4.02) | <0.01 | 90.2 | <0.01 |
| Education | No education | 5 | 3126 | 1.94 (0.91-1.83) | <0.01 | 77.6 | <0.01 |
| Occupation | No occupation | 4 | 2167 | 1.60 (0.94-2.74) | 0.08 | 81.8 | <0.01 |
| Wealth quintile | Poor | 5 | 3121 | 1.95 (1.18-3.23) | <0.01 | 82.2 | <0.01 |
| Attitude | Negative | 5 | 1845 | 1.05 (0.36-3.08) | 0.92 | 94.9 | <0.01 |
| Knowledge | Poor knowledge | 4 | 1355 | 3.23 (1.57-6.62) | <0.01 | 72.6 | 0.012 |
| Marital union | Not in a marital union | 6 | 2806 | 3.32 (1.59-6.90) | <0.01 | 89.7 | <0.01 |
| Nearest health facility distance | Far | 2 | 1156 | 1.40 (0.18-10.77) | 0.75 | 96.3 | <0.01 |

### Narrative synthesis of studies assessing Modern Contraception Use among Persons with Disabilities in Low- and Middle-Income Countries

Of the seven studies included in the narrative synthesis, we found that overall prevalence of modern contraception use was 11% to 38.1% (Table 3). These studies also reported similar list of factors, as we presented above for the studies included in the meta-analysis, associated with modern contraception use among persons with disability.

**Table 3:**
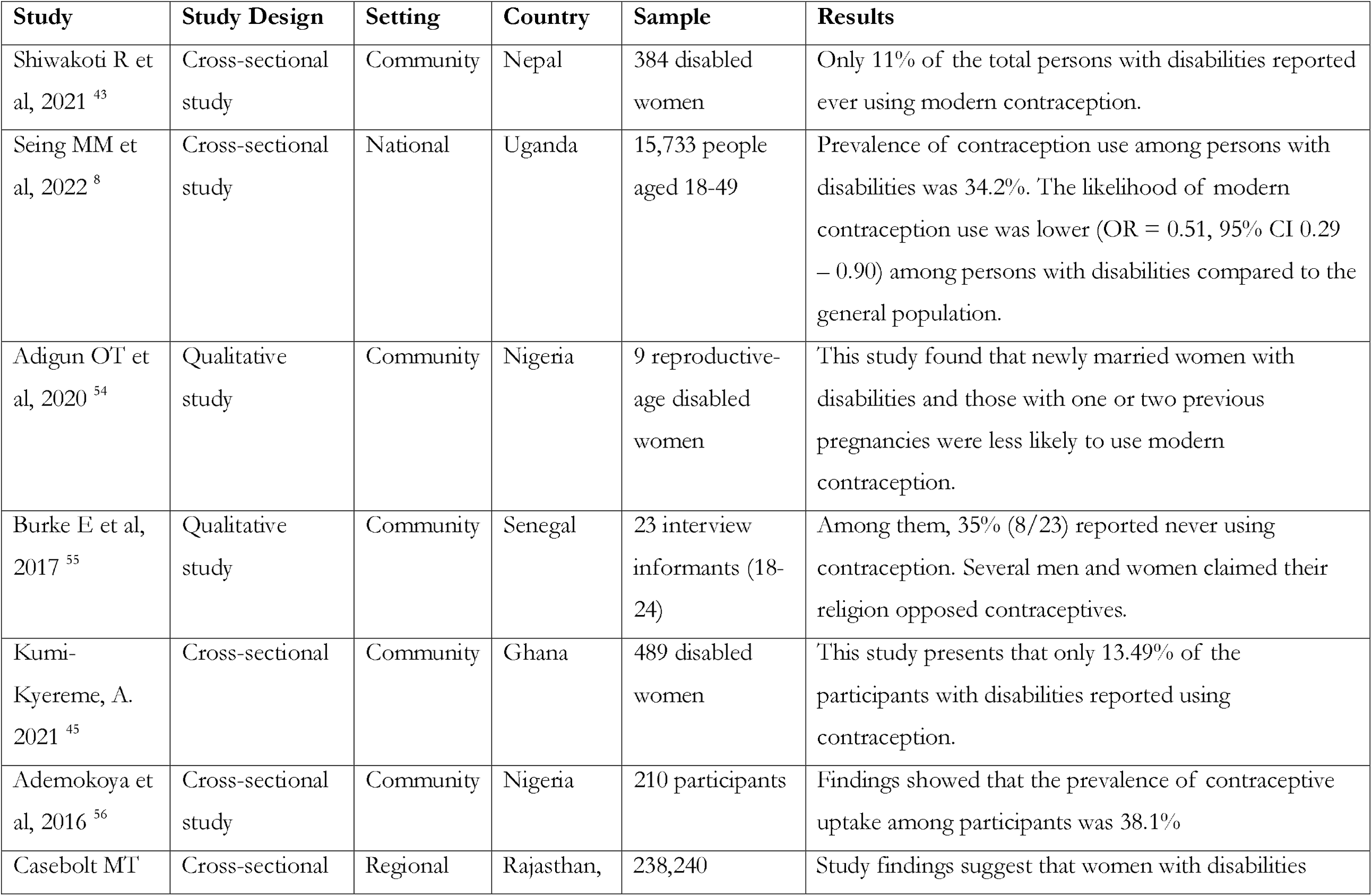
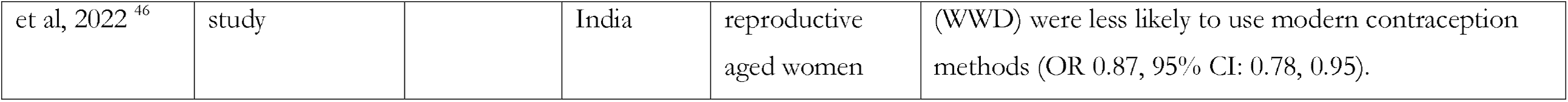
Narrative synthesis of the studies assessing contraception use among persons with disabilities in low- and lower-middle-income countries, January 2015 to June 2024.

## Discussion

The aims of this study were to explore the prevalence of modern contraception use among persons with disabilities in LMICs and identify key explanatory factors. The overall prevalence of modern contraception use was found to be 31.40% (95% CI: 26.56, 36.23), with significant heterogeneity across respondent characteristics. The highest prevalence was reported in studies with sample sizes of 5,352 or more (40.61%, 95% CI: 24.15-57.08), national-level studies (35.77%, 95% CI: 22.33-49.20), and among persons with disabilities in Bangladesh (49.00%, 95% CI: 46.25-51.75). Five factors were significantly associated with modern contraception use among persons with disabilities: age over 25 years, having some education, being in a wealth quintile other than poor, having adequate knowledge, and being in a formal marital relationship. A narrative synthesis of an additional seven studies also supports the pooled prevalence of modern contraception use. Together, these findings indicate a lower prevalence of modern contraception use among persons with disabilities in LMICs, highlighting the need for policies and programs to increase modern contraception uptake among this population to improve maternal and child health outcomes.

We found that less than one-third of persons with disabilities use contraception, which is significantly lower than the overall contraception use rate of over 60% in LMICs ^35, 36^. This disparity highlights the substantial challenges that persons with disabilities face in accessing and utilizing modern contraceptive services. The lower modern contraception use among this group is linked to several hierarchical levels of challenges ^9, 37^. First, social stigma and religious norms often view contraception as sinful or against religious beliefs ^38, 39^. Although these perceptions also affect the general population, persons with disabilities experience a greater impact due to their limited community engagement and lower exposure to family planning messages ^10, 40^ . Second, healthcare facilities and family planning workers tend to prioritize the general population, often neglecting persons with disabilities due to misconceptions about their sexual and reproductive needs ^10, 41^. This is in additional to the country level overall prevalence of modern contraception use as this study findings found lower use of modern in the country where the average use of modern contraception is also low and higher total fertility rate. Moreover, the need for privacy in discussing and accessing modern contraception, combined with the fact that persons with disabilities often stay indoors, complicates efforts to identify and engage them in family planning services ^31, 42^. In addition to the lower modern contraception use among persons with disabilities and its explanations, this study reveals another concerning issue. Our broad search identified studies from only seven LMICs, with quantitative findings reported by just four of them ^43-46^. These studies were based on small sample sizes and a limited number of confounders ^43-45^. These scenarios suggest that modern contraception issues among persons with disabilities in LMICs remain underexplored in existing research, despite the high priority given to sexual and reproductive health for the general population in these countries in line with the ongoing SDGs target.

Another major factor contributing to the lower modern contraception use among persons with disabilities is their socio-economic and demographic disadvantages. The study highlights that factor such as older age, having some level of education, being in a higher wealth quintile, possessing adequate knowledge about family planning, and being in a formal marital relationship are associated with a higher likelihood of using modern contraception among persons with disabilities ^9, 13, 27, 30-32, 34, 43^. Higher likelihoods of modern contraception use among persons with disability with higher age and being in formal relationship are found to be linked with interconnected factors ^28, 37^. Older individuals typically have greater life experience and may be more informed about family planning options, leading to a higher likelihood of modern contraceptive use ^47^. Additionally, as people age, they often face increased health risks associated with pregnancy, which can make the use of modern contraception more appealing ^48, 49^. Moreover, it is also possible that they have desired number of children with increasing of age which result in increased modern contraception use. Being in a formal relationship, such as a marriage, often provides a more stable environment for discussing and implementing family planning methods ^13, 27, 32, 34^. In such relationships, there is generally greater communication about reproductive health and shared responsibility for contraception. Moreover, formal relationships may offer better access to healthcare resources and support networks, facilitating easier access to contraceptive services.

The study identified three key factors associated with increased modern contraceptive use among individuals with disabilities: having some level of education, belonging to a higher wealth quintile, and possessing adequate knowledge about family planning ^13, 27, 30, 32, 33^. These factors often co-occur, creating a set of advantageous conditions that collectively enhance access to and use of modern contraceptive methods. Education empowers individuals with knowledge about sexual and reproductive health, including modern contraception methods, their benefits, and potential side effects ^50^. It also improves decision-making abilities, allowing individuals to make informed choices about their reproductive health. Moreover, education is often linked to better access to healthcare services and information, which facilitates easier access to contraceptive methods ^50, 51^. Similarly, individuals with disabilities in higher wealth quintiles have increased purchasing power for modern contraception, better access to healthcare services, and greater exposure to mass media about modern contraception, all of which contribute to higher modern contraceptive use in this group.

Moreover, persons with disabilities often face physical and communication barriers when accessing healthcare facilities, including family planning services. Limited accessibility of these facilities, inadequate training of healthcare providers on disability needs, and a lack of disability-friendly information and communication methods can hinder contraceptive uptake ^52, 53^. Future policies should consider improving physical access to health services, training healthcare workers on disability inclusion, and developing accessible communication tools to ensure equitable access to contraception for persons with disabilities.

This study has several strengths and a few limitations. To our knowledge, it is the first study in LMICs to summarize the prevalence of modern contraceptive use and its major contributors among individuals with disabilities. We included all studies conducted in LMICs since 2015, when the SDGs were established, regardless of their methodologies. We employed comprehensive search techniques for data extraction and the selection of eligible studies, adhering to the “Strengthening the Reporting of Observational Studies in Epidemiology” (STROBE) and PRISMA guidelines for reporting our findings. Appropriate statistical methods were utilized to provide summary estimates, enabling us to deliver more precise data for evidence-based policy and program development. However, the summary estimates presented in this study are primarily based on cross-sectional studies. We did not search for unpublished papers or grey literature, which may contribute to publication bias. Additionally, the lack of studies from a wide range of LMICs and the small sample sizes of the included studies, characterized by higher heterogeneity, limit the generalizability of our results at the national level. With two-thirds of the reviewed papers originating from Ethiopia, there is also a risk of estimation bias.

## Conclusion

This study highlights the significantly lower prevalence of modern contraception use among persons with disabilities in LMICs, with only 31.40% using modern contraception. Factors such as age, education, wealth status, knowledge of family planning, and being in a formal marital relationship are associated with modern contraception use. To increase modern contraception use among persons with disabilities in LMICs, it is crucial to improve education, address social norms, and strengthen healthcare facilities to enhance access to modern contraceptive services.

## Supporting information

Linked

linked

## Data Availability

All data produced in the present work are contained in the manuscript

## Declarations

### Data sharing statement

All relevant data are presented in the supplementary files, table and figures.

## Acknowledgement

We acknowledge the support of Department of Population science, Jatiya Kabi Kazi Nazrul Islam University, Bangladesh, where the study was conducted.

## Authors’ contributors

MNK and ARC designed the study concept. ARC conducted the formal analysis with the help of MNK. ARC, MNK, and SJK drafted the manuscript. MZI and GK reviewed the first manuscript. MNK supervised all works. All authors critically reviewed and approved the final version of this manuscript.

