## Supplementary material for "Prevalence and determinants of modern contraception use among persons with disabilities in low- and middle-income countries: a systematic review and meta-analysis": linked

**Supplementary Table 01:** Search strategy of Medline database (date: January 2015 to June 28, 2024)

| **#** | **Searches** | **Results** |
| --- | --- | --- |
| 1 | (Women or men or reproductive age).mp. [mp=title, book title, abstract, original title, name of substance word, subject heading word, floating sub-heading word, keyword heading word, organism supplementary concept word, protocol supplementary concept word, rare disease supplementary concept word, unique identifier, synonyms, population supplementary concept word, anatomy supplementary concept word] | 1225197 |
| 2 | (Disability* or disability* or impairment* or physical disability* or visual impairment* or visual loss or blind or blind or hearing loss or hearing loss or hearing impairment* or deaf or intellectual disability* or intellectual disability* or sensory disability).mp. [mp=title, book title, abstract, original title, name of substance word, subject heading word, floating sub-heading word, keyword heading word, organism supplementary concept word, protocol supplementary concept word, rare disease supplementary concept word, unique identifier, synonyms, population supplementary concept word, anatomy supplementary concept word] | 1143702 |
| 3 | (Birth control or family planning service* or family planning services or contraception behavior or modern contraception* or traditional contraception* or barrier* or facilitator*).mp. [mp=title, book title, abstract, original title, name of substance word, subject heading word, floating sub-heading word, keyword heading word, organism supplementary concept word, protocol supplementary concept word, rare disease supplementary concept word, unique identifier, synonyms, population supplementary concept word, anatomy supplementary concept word] | 500541 |
| 4 | (Afghani* or Guinea* or Rwanda* or Benin* or Guinea-Bissau* or Senegal* or Burkina Faso* or Haiti* or Sierra Leone* or Burundi* or Korea* or Somalia* or Central African Republic or Liberia* or South Sudan* or Chad* or Madagascar* or Tanzania* or Comoros* or Malawi* or Togo* or Congo* or Mali* or Uganda* or Eritrea* or Mozambique* or Zimbabwe* or Ethiopia* or Nepa* or Gambia* or Niger* or Angol* or Indonesia* or Philippine* or Armenia* or Jordan* or Sao Tome* or Bangladesh* or Kenya* or Solomon Island* or Bhutan* or Kiribati* or Sri Lanka* or Bolivia* or Kosovo* or Sudan* or Cabo Verde* or Kyrgyz Republic* or Swazi* or Cambodia* or Lao* or Syria* or Cameroon* or Lesotho* or Tajikistan* or Congo* or Mauritania* or Timor-Leste* or Cote d'Ivoire* or Micronesia* or Tunisia* or Djibouti* or Moldov* or Ukraine* or Egypt* or Mongolia* or Uzbekistan* or El Salvador* or Morocco* or Vanuat* or Georgia* or Myanmar* or Vietnam* or Ghana* or Nicaragua* or West Bank* or Gaza* or Guatemala* or Nigeria* or Yemen* or Honduras* or Pakistan* or Zambia* or India* or Papua New Guinea*).mp. [mp=title, book title, abstract, original title, name of substance word, subject heading word, floating sub-heading word, keyword heading word, organism supplementary concept word, protocol supplementary concept word, rare disease supplementary concept word, unique identifier, synonyms, population supplementary concept word, anatomy supplementary concept word] | 1920085 |
| 5 | (((low* or middle) adj income adj (country* or nation*)) or LMICs).mp. [mp=title, book title, abstract, original title, name of substance word, subject heading word, floating sub-heading word, keyword heading word, organism supplementary concept word, protocol supplementary concept word, rare disease supplementary concept word, unique identifier, synonyms, population supplementary concept word, anatomy supplementary concept word] | 49006 |
| 6 | 1 and 2 | 65822 |
| 7 | 4 or 5 | 1949323 |
| 8 | 3 and 6 and 7 | 238 |
| 9 | limit 8 to yr="2015 -Current" | 157 |

**Supplementary Table 02:** Search strategy of PsycInfo  database (date: January 2015 to June 28, 2024)

| **#** | **Searches** | **Results** |
| --- | --- | --- |
| 1 | (Women or women or reproductive age).mp. [mp=title, abstract, heading word, table of contents, key concepts, original title, tests & measures, mesh word] | 328141 |
| 2 | (Disability* or disability* or impairment* or physical disability* or visual impairment* or visual loss or blind or blind or hearing loss or hearing loss or hearing impairment* or deaf or intellectual disability* or intellectual disability* or sensory disability).mp. [mp=title, abstract, heading word, table of contents, key concepts, original title, tests & measures, mesh word] | 422670 |
| 3 | (Birth control or family planning service* or family planning services or contraception behavior or modern contraception* or traditional contraception* or barrier* or facilitator*).mp. [mp=title, abstract, heading word, table of contents, key concepts, original title, tests & measures, mesh word] | 122631 |
| 4 | (Afghani* or Guinea* or Rwanda* or Benin* or Guinea-Bissau* or Senegal* or Burkina Faso* or Haiti* or Sierra Leone* or Burundi* or Korea* or Somalia* or Central African Republic or Liberia* or South Sudan* or Chad* or Madagascar* or Tanzania* or Comoros* or Malawi* or Togo* or Congo* or Mali* or Uganda* or Eritrea* or Mozambique* or Zimbabwe* or Ethiopia* or Nepa* or Gambia* or Niger* or Angol* or Indonesia* or Philippine* or Armenia* or Jordan* or Sao Tome* or Bangladesh* or Kenya* or Solomon Island* or Bhutan* or Kiribati* or Sri Lanka* or Bolivia* or Kosovo* or Sudan* or Cabo Verde* or Kyrgyz Republic* or Swazi* or Cambodia* or Lao* or Syria* or Cameroon* or Lesotho* or Tajikistan* or Congo* or Mauritania* or Timor-Leste* or Cote d'Ivoire* or Micronesia* or Tunisia* or Djibouti* or Moldov* or Ukraine* or Egypt* or Mongolia* or Uzbekistan* or El Salvador* or Morocco* or Vanuat* or Georgia* or Myanmar* or Vietnam* or Ghana* or Nicaragua* or West Bank* or Gaza* or Guatemala* or Nigeria* or Yemen* or Honduras* or Pakistan* or Zambia* or India* or Papua New Guinea*).mp. [mp=title, abstract, heading word, table of contents, key concepts, original title, tests & measures, mesh word] | 188863 |
| 5 | (((low* or middle) adj income adj (countr* or nation*)) or LMICs).mp. [mp=title, abstract, heading word, table of contents, key concepts, original title, tests & measures, mesh word] | 7884 |
| 6 | 1 and 2 | 18503 |
| 7 | 4 or 5 | 193588 |
| 8 | 3 and 6 and 7 | 85 |
| 9 | limit 8 to yr="2015 -Current" | 55 |

**Supplementary Table 03:** Search strategy of Embase database (date: January 2015 to June 28, 2024)

| **#** | **Searches** | **Results** |
| --- | --- | --- |
| 1 | (Women or women or reproductive age).mp. [mp=title, abstract, heading word, drug trade name, original title, device manufacturer, drug manufacturer, device trade name, keyword heading word, floating subheading word, candidate term word] | 1741004 |
| 2 | (Disabili* or disabili* or impairmen* or physical disabilit* or visual impairmen* or visual loss or blind or blind or hearing loss or hearing loss or hearing impairmen* or deaf or intellectual disabili* or intellectual disabili* or sensory disabili).mp. [mp=title, abstract, heading word, drug trade name, original title, device manufacturer, drug manufacturer, device trade name, keyword heading word, floating subheading word, candidate term word] | 1666623 |
| 3 | (Birth control or family planning service* or family planning services or contraception behavior or modern contracepti* or traditional contracepti* or barrier* or facilitator*).mp. [mp=title, abstract, heading word, drug trade name, original title, device manufacturer, drug manufacturer, device trade name, keyword heading word, floating subheading word, candidate term word] | 627096 |
| 4 | (Afghani* or Guinea* or Rwand* or Benin* or Guinea-Bissau* or Senegal* or Burkina Faso* or Haiti* or Sierra Leone* or Burundi* or Korea* or Somalia* or Central African Republic or Liberia* or South Sudan* or Chad* or Madagasca* or Tanzania* or Comoros* or Malawi* or Togo* or Congo* or Mali* or Uganda* or Eritrea* or Mozambique* or Zimbabwe* or Ethiopia* or Nepa* or Gambia* or Niger* or Angol* or Indonesia* or Philippin* or Armenia* or Jordan* or Sao Tome* or Bangladesh* or Kenya* or Solomon Island* or Bhutan* or Kiribati* or Sri Lanka* or Bolivia* or Kosov* or Sudan* or Cabo Verde* or Kyrgyz Republic* or Swazi* or Cambodia* or Lao* or Syria* or Cameroon* or Lesotho* or Tajikistan* or Congo* or Mauritania* or Timor-Leste* or Cote d'Ivoire* or Micronesia* or Tunisia* or Djibouti* or Moldov* or Ukraine* or Egypt* or Mongolia* or Uzbekistan* or El Salvador* or Morocco* or Vanuat* or Georgia* or Myanmar* or Vietnam* or Ghana* or Nicaragua* or West Bank* or Gaza* or Guatemala* or Nigeria* or Yemen* or Hondur* or Pakistan* or Zambia* or India* or Papua New Guinea*).mp. [mp=title, abstract, heading word, drug trade name, original title, device manufacturer, drug manufacturer, device trade name, keyword heading word, floating subheading word, candidate term word] | 2857750 |
| 5 | (((low* or middle) adj income adj (countr* or nation*)) or LMICs).mp. [mp=title, abstract, heading word, drug trade name, original title, device manufacturer, drug manufacturer, device trade name, keyword heading word, floating subheading word, candidate term word] | 66180 |
| 6 | 1 and 2 | 101392 |
| 7 | 4 or 5 | 2897330 |
| 8 | 3 and 6 and 7 | 296 |
| 9 | limit 8 to yr="2015 -Current" | 212 |

**Supplementary Table 04:** Search strategy of Emcare database (date: January 2015 to June 28, 2024)

| **#** | **Searches** | **Results** |
| --- | --- | --- |
| 1 | (Women or women or reproductive age).mp. [mp=title, abstract, heading word, drug trade name, original title, device manufacturer, drug manufacturer, device trade name, keyword heading word] | 554356 |
| 2 | (Disabili* or disabili* or impairmen* or physical disabilit* or visual impairmen* or visual loss or blind or blind or hearing loss or hearing loss or hearing impairmen* or deaf or intellectual disabili* or intellectual disabili* or sensory disabili).mp. [mp=title, abstract, heading word, drug trade name, original title, device manufacturer, drug manufacturer, device trade name, keyword heading word] | 454959 |
| 3 | (Birth control or family planning service* or family planning services or contraception behavior or modern contracepti* or traditional contracepti* or barrier* or facilitator*).mp. [mp=title, abstract, heading word, drug trade name, original title, device manufacturer, drug manufacturer, device trade name, keyword heading word] | 177453 |
| 4 | (Afghani* or Guinea* or Rwand* or Benin* or Guinea-Bissau* or Senegal* or Burkina Faso* or Haiti* or Sierra Leone* or Burundi* or Korea* or Somalia* or Central African Republic or Liberia* or South Sudan* or Chad* or Madagasca* or Tanzania* or Comoros* or Malawi* or Togo* or Congo* or Mali* or Uganda* or Eritrea* or Mozambique* or Zimbabwe* or Ethiopia* or Nepa* or Gambia* or Niger* or Angol* or Indonesia* or Philippin* or Armenia* or Jordan* or Sao Tome* or Bangladesh* or Kenya* or Solomon Island* or Bhutan* or Kiribati* or Sri Lanka* or Bolivia* or Kosov* or Sudan* or Cabo Verde* or Kyrgyz Republic* or Swazi* or Cambodia* or Lao* or Syria* or Cameroon* or Lesotho* or Tajikistan* or Congo* or Mauritania* or Timor-Leste* or Cote d'Ivoire* or Micronesia* or Tunisia* or Djibouti* or Moldov* or Ukrain* or Egypt* or Mongolia* or Uzbekistan* or El Salvador* or Morocc* or Vanuat* or Georgia* or Myanmar* or Vietnam* or Ghana* or Nicaragua* or West Bank* or Gaza* or Guatemala* or Nigeria* or Yemen* or Hondur* or Pakistan* or Zambia* or India* or Papua New Guinea*).mp. [mp=title, abstract, heading word, drug trade name, original title, device manufacturer, drug manufacturer, device trade name, keyword heading word] | 506638 |
| 5 | (((low* or middle) adj income adj (countr* or nation*)) or LMICs).mp. [mp=title, abstract, heading word, drug trade name, original title, device manufacturer, drug manufacturer, device trade name, keyword heading word] | 27826 |
| 6 | 1 and 2 | 34015 |
| 7 | 4 or 5 | 522882 |
| 8 | 3 and 6 and 7 | 142 |
| 9 | limit 8 to yr="2015 -Current" | 104 |

**Supplementary Table 05:** Search strategy of Maternity and Infant Care  database (date: January 2015 to June 28, 2024)

| **#** | **Searches** | **Results** |
| --- | --- | --- |
| 1 | (Women or women or reproductive age).mp. [mp=abstract, heading word, title] | 123795 |
| 2 | (Disabili* or disabili* or impairmen* or physical disabilit* or visual impairmen* or visual loss or blind or blind or hearing loss or hearing loss or hearing impairmen* or deaf or intellectual disabili* or intellectual disabili* or sensory disabili).mp. [mp=abstract, heading word, title] | 10330 |
| 3 | (Birth control or family planning service* or family planning services or contraception behavior or modern contracepti* or traditional contracepti* or barrier* or facilitator*).mp. [mp=abstract, heading word, title] | 8351 |
| 4 | (Afghani* or Guinea* or Rwand* or Benin* or Guinea-Bissau* or Senegal* or Burkina Faso* or Haiti* or Sierra Leone* or Burundi* or Korea* or Somalia* or Central African Republic or Liberia* or South Sudan* or Chad* or Madagasca* or Tanzania* or Comoros* or Malawi* or Togo* or Congo* or Mali* or Uganda* or Eritrea* or Mozambique* or Zimbabwe* or Ethiopia* or Nepa* or Gambia* or Niger* or Angol* or Indonesia* or Philippin* or Armenia* or Jordan* or Sao Tome* or Bangladesh* or Kenya* or Solomon Island* or Bhutan* or Kiribati* or Sri Lanka* or Bolivia* or Kosov* or Sudan* or Cabo Verde* or Kyrgyz Republic* or Swazi* or Cambodia* or Lao* or Syria* or Cameroon* or Lesotho* or Tajikistan* or Congo* or Mauritania* or Timor-Leste* or Cote d'Ivoire* or Micronesia* or Tunisia* or Djibouti* or Moldov* or Ukrain* or Egypt* or Mongolia* or Uzbekistan* or El Salvador* or Morocc* or Vanuat* or Georgia* or Myanmar* or Vietnam* or Ghana* or Nicaragua* or West Bank* or Gaza* or Guatemala* or Nigeria* or Yemen* or Hondur* or Pakistan* or Zambia* or India* or Papua New Guinea*).mp. [mp=abstract, heading word, title] | 23392 |
| 5 | (((low* or middle) adj income adj (countr* or nation*)) or LMICs).mp. [mp=abstract, heading word, title] | 2698 |
| 6 | 1 and 2 | 3293 |
| 7 | 4 or 5 | 24901 |
| 8 | 3 and 6 and 7 | 14 |
| 9 | limit 8 to yr="2015 -Current" | 11 |

**Supplementary Table 06:** Search strategy of Scopus  database (date: January 2015 to June 28, 2024)

| Searches | Results |
| --- | --- |
| ( ( TITLE-ABS- ( women OR men OR reproductive AND age ) AND ( disabili* OR disabili* OR impairmen* OR physical AND disabilit* OR visual AND impairmen* OR visual AND loss OR blind OR blind OR hearing AND loss OR hearing AND loss OR hearing AND impairmen* OR deaf OR intellectual AND disabili* OR intellectual AND disabili* OR sensory AND disabili ) AND  ( ( TITLE-ABS-KEY ( birth AND control OR family AND planning AND service* OR family AND planning AND services OR contraception AND behavior OR modern AND contracepti* OR traditional AND contracepti* OR barrier* OR facilitator* ) AND TITLE-ABS-KEY ( eritrea*  OR  mozambique*  OR  zimbabwe*  OR  ethiopia*  OR  nepa*  OR  gambia*  OR  niger*  OR  angol*  OR  indonesia*  OR  philippin*  OR  armenia*  OR  jordan*  OR  sao  AND tome*  OR  bangladesh*  OR  kenya*  OR  solomon  AND island*  OR  bhutan*  OR  kiribati*  OR  sri  AND lanka*  OR  bolivia* )  OR  TITLE-ABS-KEY ( kosov*  OR  sudan*  OR  cabo  AND verde*  OR  kyrgyz  AND republic*  OR  swazi*  OR  cambodia*  OR  lao*  OR  syria*  OR  cameroon*  OR  lesotho*  OR  tajikistan*  OR  congo*  OR  mauritania*  OR  timor-leste*  OR  cote  AND d'ivoire*  OR  micronesia*  OR  tunisia*  OR  djibouti* )  OR  TITLE-ABS-KEY ( moldov*  OR  ukrain*  OR  egypt*  OR  mongolia*  OR  uzbekistan*  OR  el  AND salvador*  OR  morocc*  OR  vanuat*  OR  georgia*  OR  myanmar*  OR  vietnam*  OR  ghana*  OR  nicaragua*  OR  west  AND bank*  OR  gaza*  OR  guatemala*  OR  nigeria*  OR  yemen* )  OR  TITLE-ABS-KEY ( hondur*  OR  pakistan*  OR  zambia*  OR  india*  OR  papua  AND new  AND guinea* ) )  AND  PUBYEAR  >  2015 ) | 25 |

**Supplemental Table 07**. Summary of the selected studies to study modern contraception use among persons with disability in low- and middle-income countries, January 2015- June 2024.

| **Author, Year** | **Country** | **Study design and settings** | **Sample** | **Outcomes** | **Confounders adjustment** |
| --- | --- | --- | --- | --- | --- |
| Tenaw et al 2023 [1] | Ethiopia | Community based,  Cross-sectional | 620 | Contraceptive Utilization | Age, education, occupation, marital union, knowledge, transport facility, distance to health facility, wealth index, residence. |
| Yesgat YM et al 2020 [2] | Ethiopia | Community based,  Cross-sectional | 418 | Utilization of Family Planning Methods | Age, education, occupation, marital union, knowledge, wealth, attitude, parity, friendly SRH services. |
| Mekonnen et al 2020 [3] | Ethiopia | Community based,  Cross-sectional | 397 | Utilization of Family Planning Methods | Married, age, knowledge. |
| Yimer AS et al 2019 [4] | Ethiopia | Facility based,  Cross-sectional | 326 | Use of contraceptive methods | Married, age, knowledge, attitude. |
| Oumer et al 2020 [5] | Ethiopia | Community based,  Cross-sectional | 708 | Modern Contraceptive Method Utilization | Age, education, occupation, marital union, wealth, husband’s education, husband’s occupation. |
| Ayiga N et al 2016 [6] | Uganda | National,  Cross-sectional | 1128 | Uptake of Contraception | Age, education, marital union, wealth, children alive, access to health facility, number of ANC visit. |
| Beyene et al 2019 [7] | Ethiopia | Community based,  Cross-sectional | 267 | Modern contraceptive use and associated factors | Age, education, marital union, wealth |
| Makau PM et al 2021 [8] | Kenya | Community based,  Cross-sectional | 291 | Utilization of family planning | Attitude towards contraception |
| Mahmood S et al 2022 [9] | Pakistan | National,  Cross-sectional | 6711 | Utilization of essential reproductive health services | Disability status |
| Alam MB et al 2024 [10] | Bangladesh | National,  Cross-sectional | 47465 | Distribution of contraceptive methods use | Disability status, disability level, six-dimension of disability |
| Tsegay K et al 2017 [11] | Ethiopia | Community based,  Cross-sectional | 539 | Modern Contraceptive Methods Utilization | Age, marriage, distance of health facility, attitude, health facility privacy, discussion with family |
| Shiwakoti et al, 2021 [12] | Nepal | Community based, cross sectional | 384 women with disabilities | Utilization of sexual and reproductive healthcare services, including modern contraception access | Age, family type, education of caregiver, marital status, household size, occupation, disability type, disability severity, empowerment, listens to radio, perceived need for SRH, perceived susceptibility to SRH-related services, perceived severity of SRH-related services, self-efficacy |
| Seing MM et al, 2022 [13] | Uganda | National, cross sectional | 15,733 persons aged 18-49 | Modern contraception uptake | Disability type, year, sex, marital status, education, religion, wealth, region, violence |
| Adigun OT et al, 2020 [14] | Nigeria | Community based qualitative approach, one to one interview with sign language | 9 reproductive age disable women | Experiences and satisfaction of pregnant deaf women with antenatal care, contraception use | Not applicable |
| Burke E et al, 2017 [15] | Senegal | qualitative peer-to-peer approach, community based | 23 interview informants aged (18-24) | Access to sexual and reproductive health services, including modern contraception use | Not applicable |
| Kumi-Kyereme, A. 2021 [16] | Ghana | Community based, cross sectional | 489 disable women | Sexual and reproductive health services use, including modern contraception use. | Not applicable |
| Ademokoya et al, 2016 [17] | Nigeria | Community based, descriptive study | 210 participants | Sexual behaviours and modern contraceptive use | Not applicable |
| Casebolt MT et al, 2022 [18] | Rajasthan, India | National, cross sectional | 238,240 women | Modern contraceptive use | Not applicable |

.

**Quality assessment of the included studies**

**Supplemental Table 08.** Newcastle-Ottawa scale assessment of study quality for **cross-sectional study**

| **Author, Year** | Selection | | | |  | Comparability |  | Outcome | | Study quality |
| --- | --- | --- | --- | --- | --- | --- | --- | --- | --- | --- |
|  | 1 | 2 | 3 | 4 |  | 5 |  | 6 | 7 |  |
|  | Representativeness of the sample | Sample size | Ascertainment of exposure | Non-respondents |  | The subjects in different outcome groups are comparable, based on the study design or analysis. Confounding factors are controlled. |  | Assessment of outcome | Statistical test is appropriate |  |
| Tenaw et al 2023 [1] | * | * | * | * |  | * |  | * | * | 7 |
| Yesgat YM et al 2020 [2] | * | * | * | * |  | * |  | * |  | 6 |
| Mekonnen et al 2020 [3] | * | * | * | * |  | * |  | * | * | 7 |
| Yimer AS et al 2019 [4] | * | * | * | * |  | * |  | * | * | 7 |
| Oumer et al 2020 [5] | * |  | * | * |  | * |  | * | * | 6 |
| Ayiga N et al 2016 [6] | * | * | * | * |  | * |  | * | * | 7 |
| Beyene et al 2019 [7] | * | * | * | * |  | * |  | * | * | 7 |
| Makau PM et al 2021 [8] | * | * | * | * |  | * |  | * | * | 7 |
| Mahmood S et al 2022 [9] | * | * | * | * |  | * |  | * | * | 7 |
| Alam MB et al 2024 [10] | * | * | * | * |  | * |  | * | * | 7 |
| Tsegay K et al 2017 [11] | * | * | * | * |  | * |  | * | * | 7 |
| Shiwakoti et al, 2021 [12] | * | * | * | * |  | * |  | * | * | 7 |
| Seing MM et al, 2022 [13] | * | * | * | * |  | * |  | * | * | 7 |
| Kumi-Kyereme, A. 2021 [16] | * | * | * | * |  | * |  | * | * | 7 |
| Ademokoya et al, 2016 [17] | * | * | * | * |  | * |  | * | * | 7 |
| Casebolt MT et al, 2022 [18] | * | * | * | * |  | * |  | * | * | 7 |

**Supplemental Table 09.** Newcastle-Ottawa scale assessment of study quality for **qualitative study**

|  | Research aim(s) was/were clearly stated | Qualitative methods are an appropriate approach to this issue | Study design was suitable for answering the research question | Recruitment strategy was appropriate for the aims of the research | Data collection was adequate for answering the research question | Relationship between researcher and participants was adequately considered | Other potential ethical issues were adequately considered | Data analysis was of sufficient rigor | Findings were clearly stated | Research adds value to science, practice, and/or policy | Study quality |
| --- | --- | --- | --- | --- | --- | --- | --- | --- | --- | --- | --- |
| Adigun OT et al, 2020 [14] | * | * | * | * | * | * | * | * | * | * | 10 |
| Burke E et al, 2017 [15] | * | * | * | * | * | * | * | * | * | * | 10 |

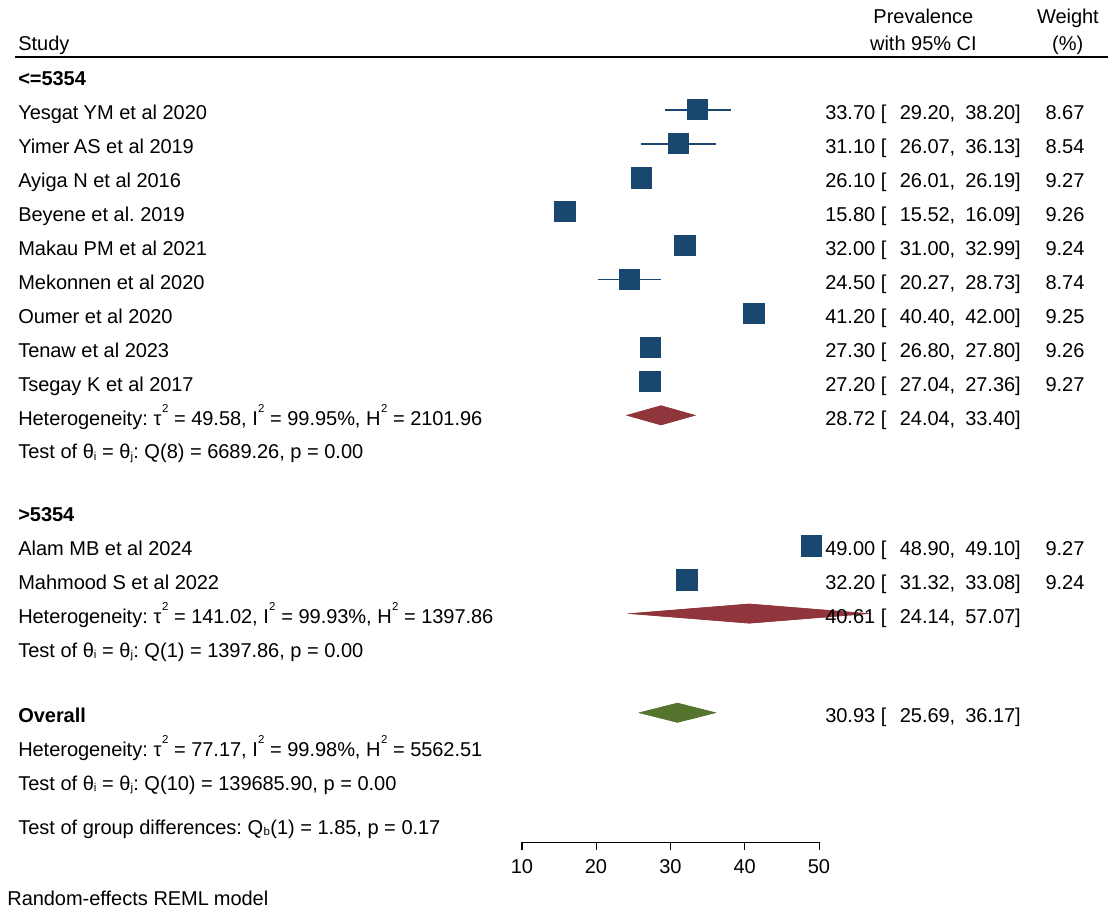

**Supplementary figure 1:** Heterogeneity of modern contraception use among persons with disability in low- and middle-income countries by sample size of the included studies

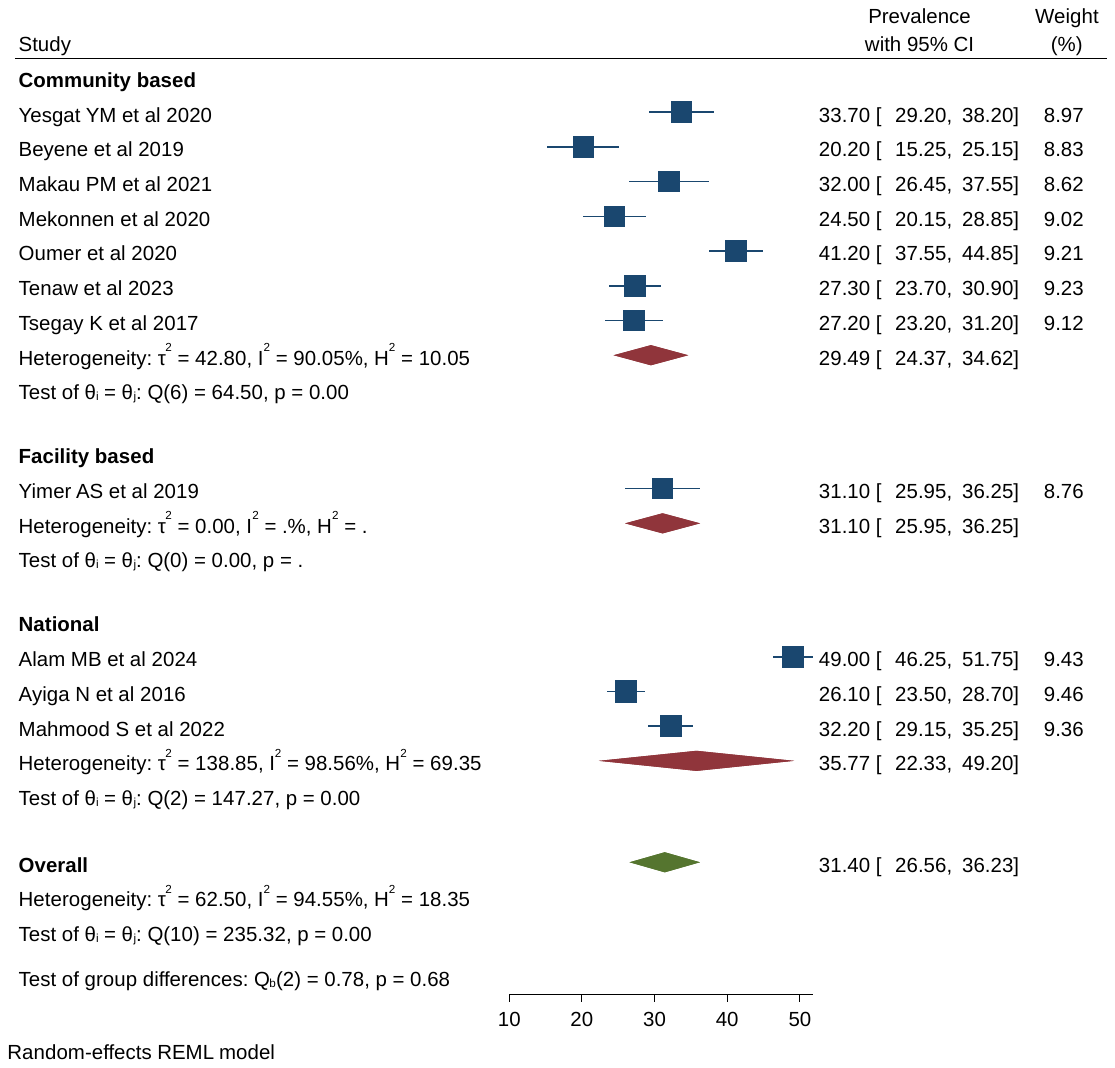

**Supplementary figure 2:** Heterogeneity of modern contraception use among persons with disability in low- and middle-income countries by study settings of the included studies

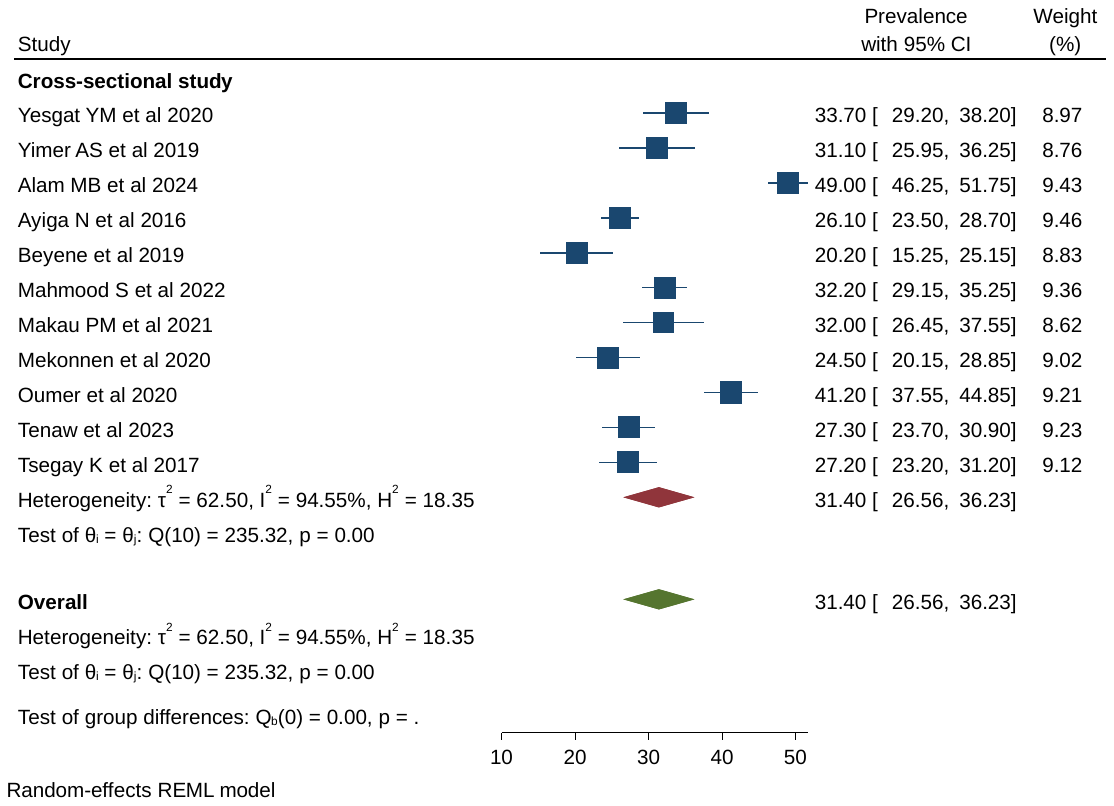

**Supplementary figure 3:** Heterogeneity of modern contraception use among persons with disability in low- and middle-income countries by study design of the included studies.

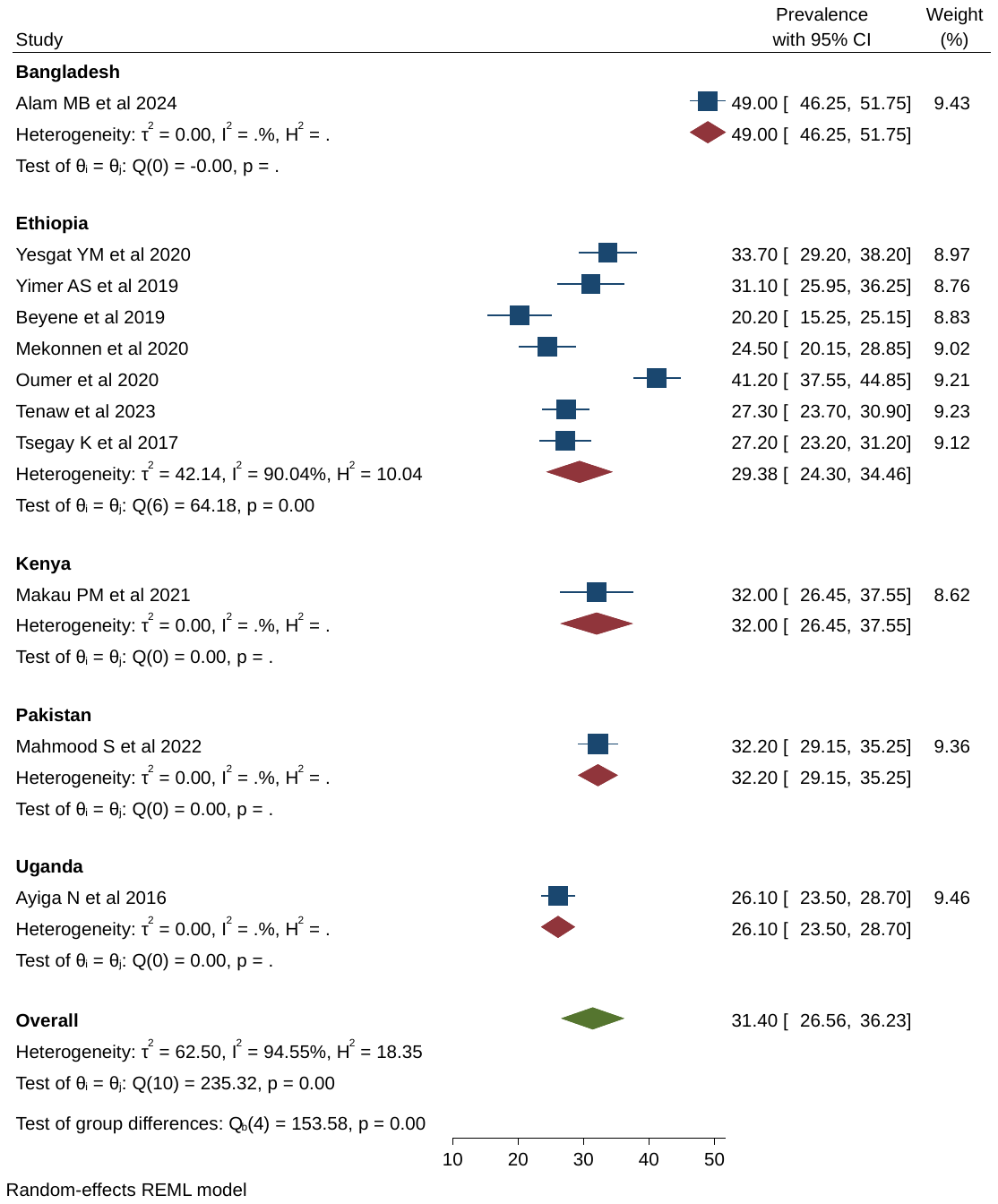

**Supplementary figure 4:** Heterogeneity of modern contraception use among persons with disability in low- and middle-income countries by country of the included studies.

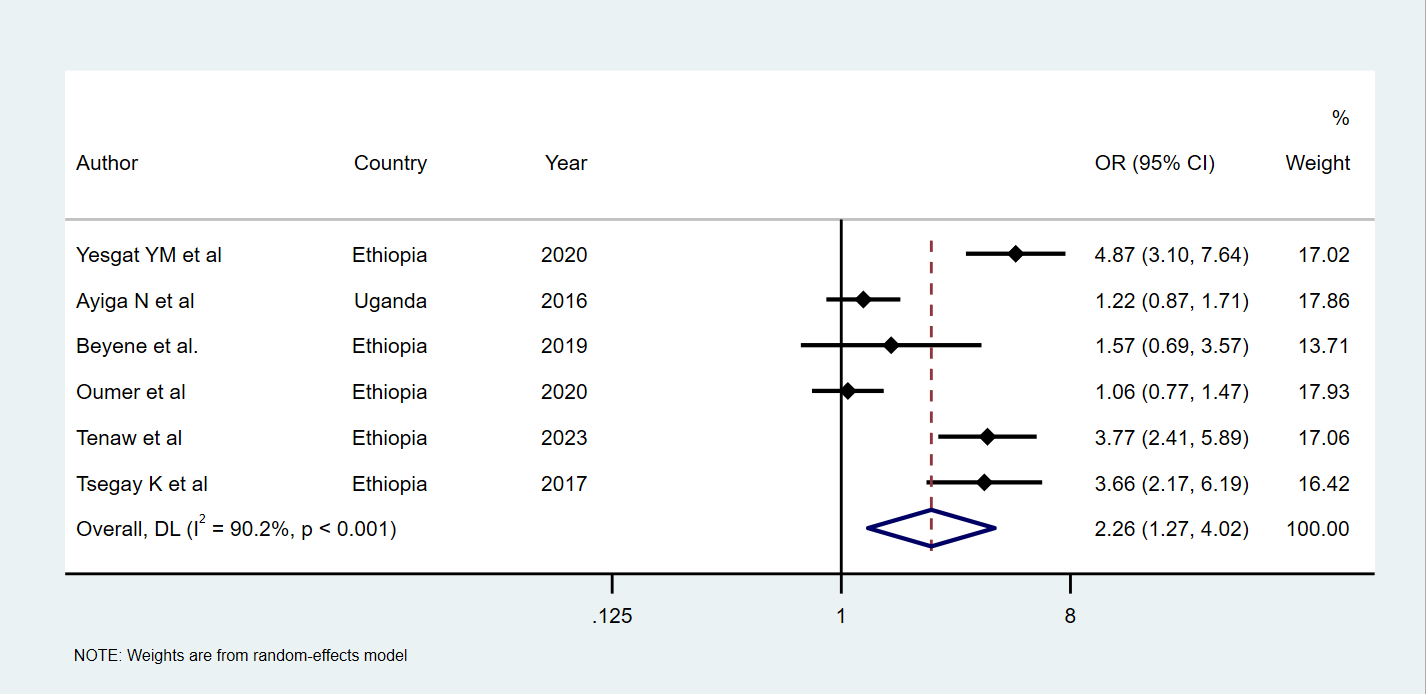

**Supplementary figure 5:** Summary estimations of age on modern contraception uptake among women with disabilities in low- and middle-income countries.

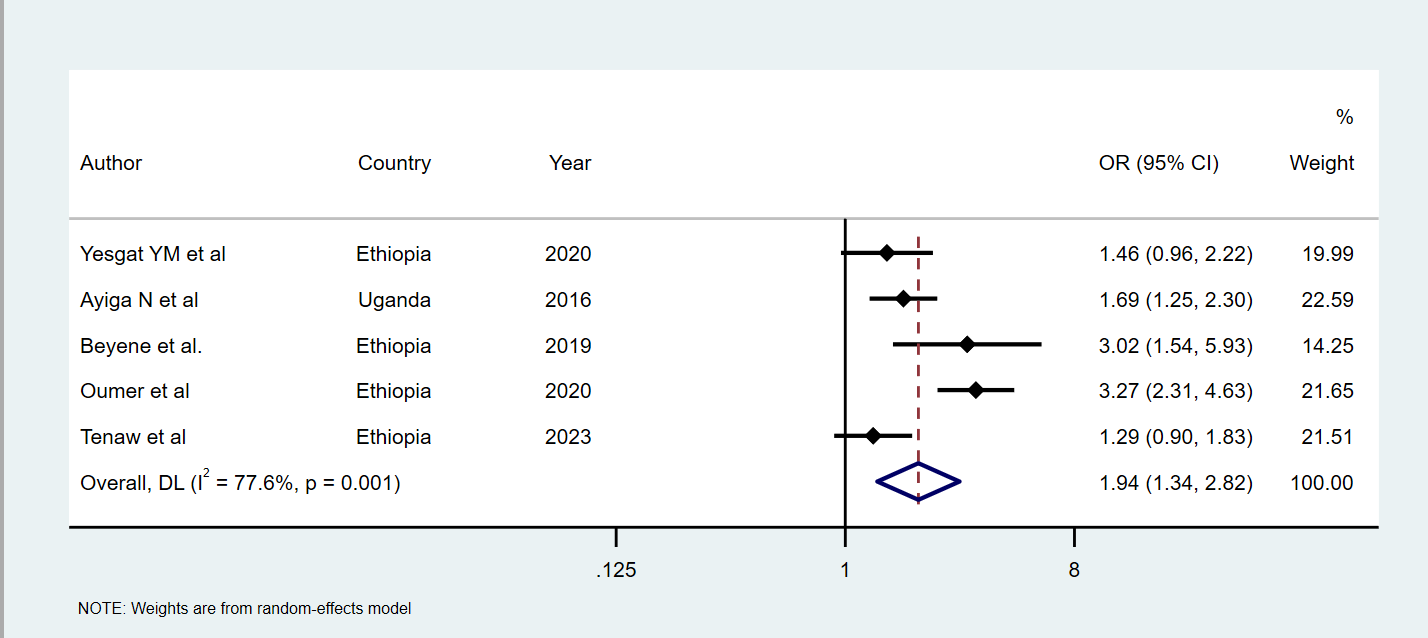

**Supplementary figure 6:** Summary estimations of education on modern contraception uptake among women with disabilities in low- and middle-income countries

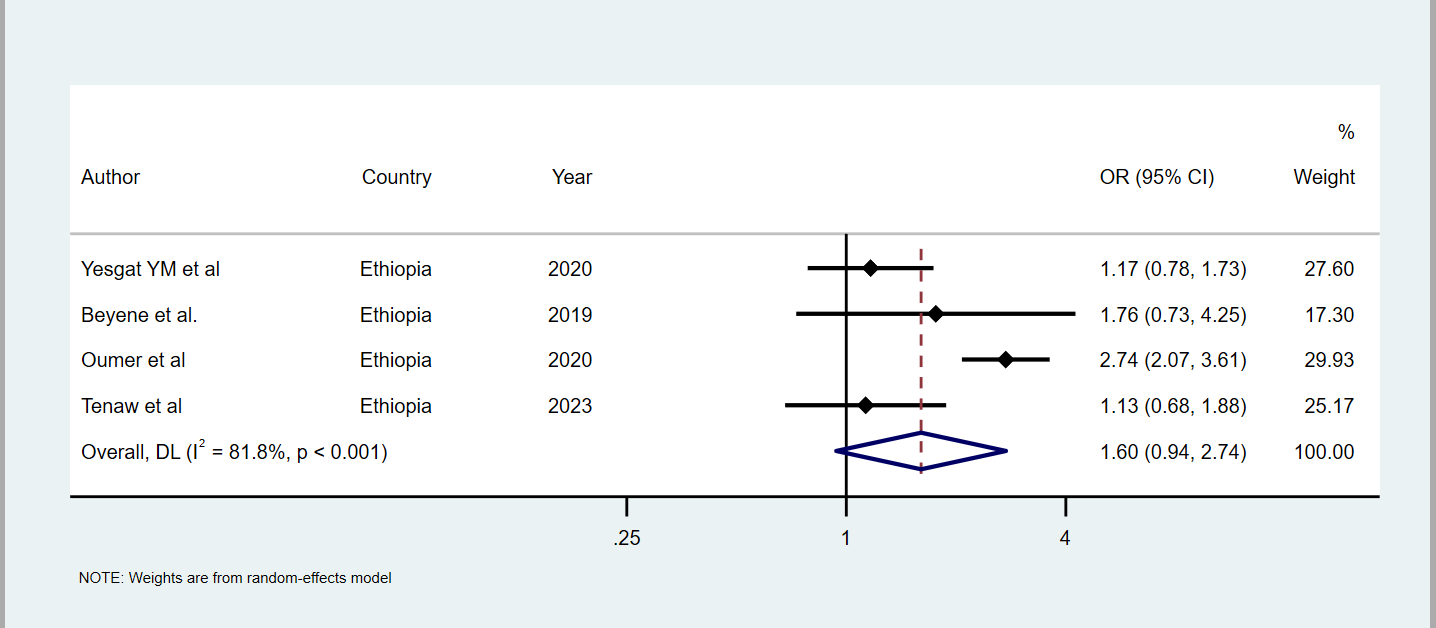

**Supplementary figure 7:** Summary estimations of occupation on modern contraception uptake among women with disabilities in low- and middle-income countries.

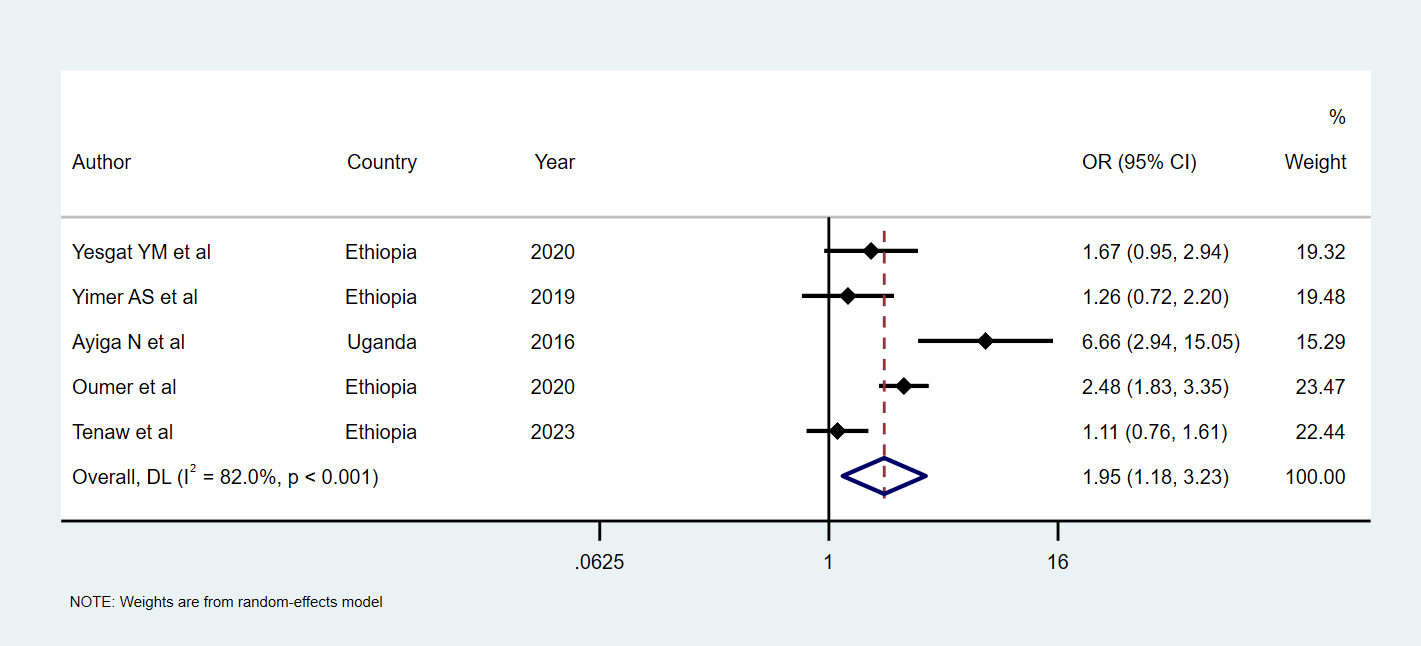

**Supplementary figure 8:** Summary estimations of wealth index on modern contraception uptake among women with disabilities in low- and middle-income countries.

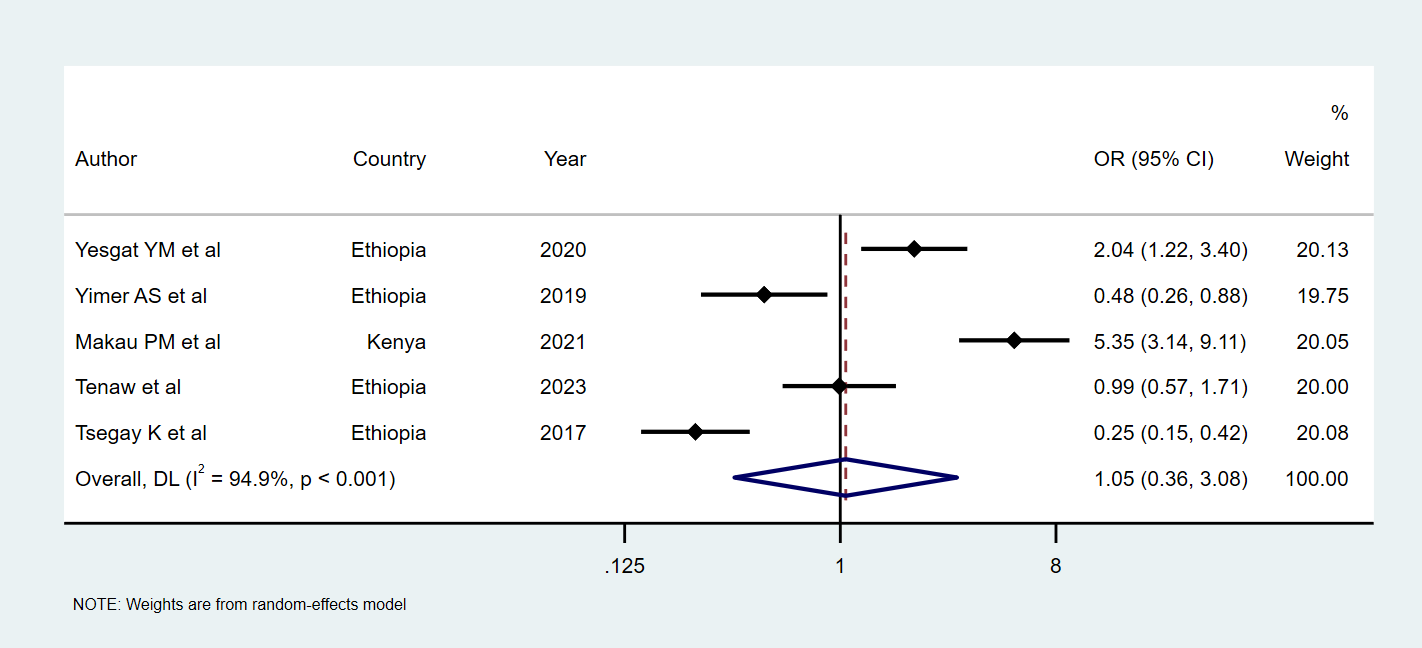

**Supplementary figure 9:** Summary estimations of attitude on modern contraception uptake among women with disabilities in low- and middle-income countries.

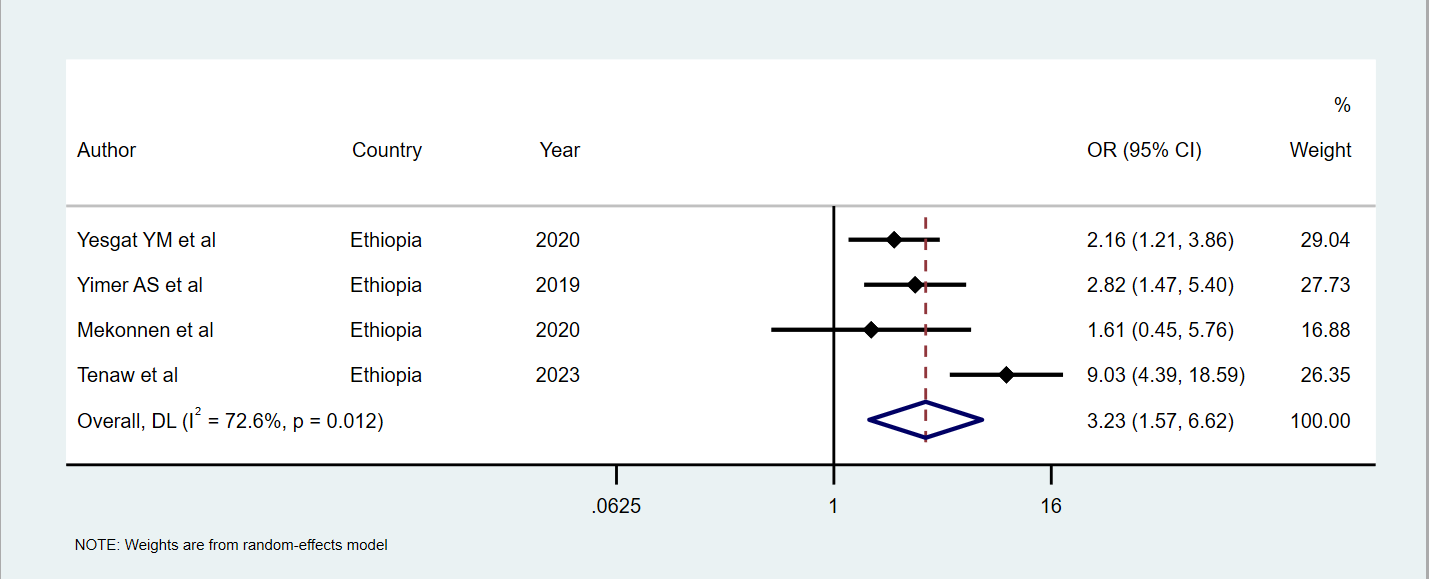

**Supplementary figure 10:** Summary estimations of knowledge on modern contraception uptake among women with disabilities in low- and middle-income countries.

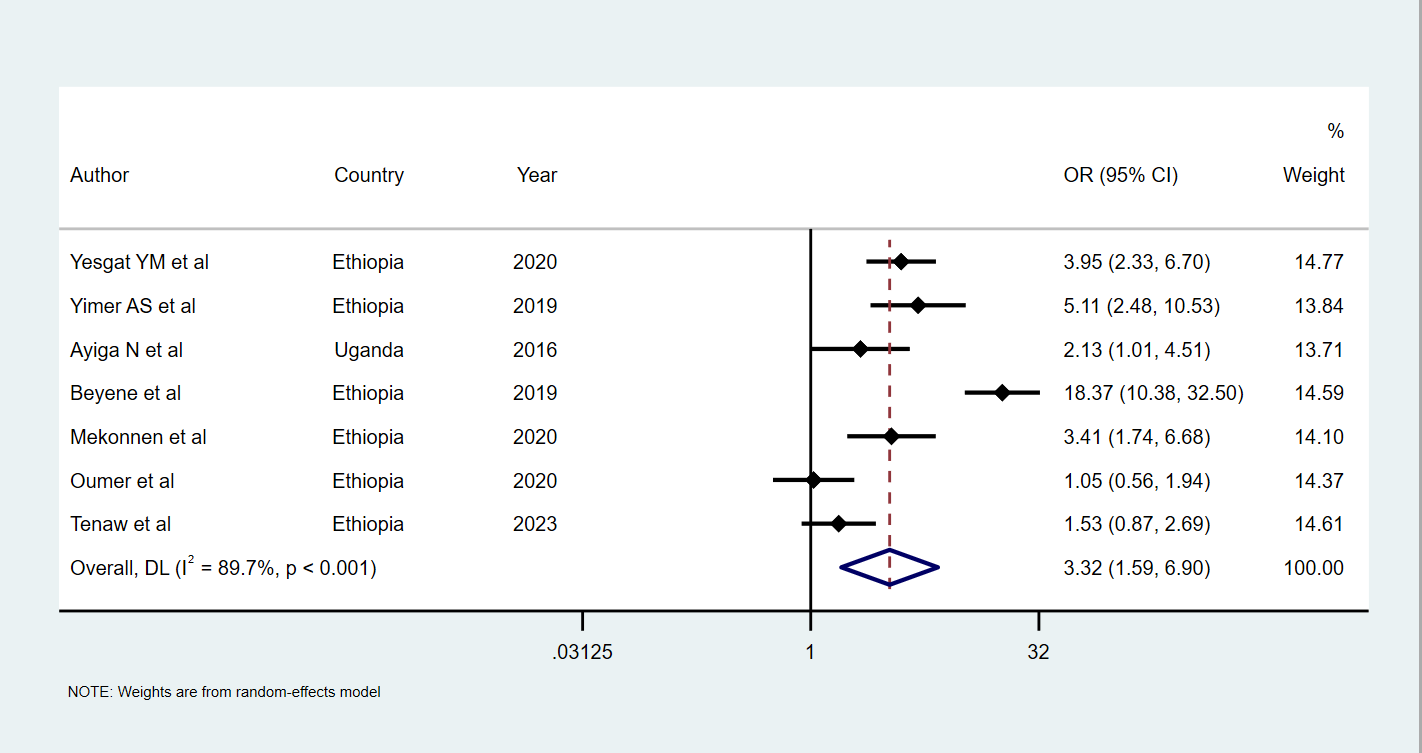

**Supplementary figure 11:** Summary estimations of marital union on modern contraception uptake among women with disabilities in low- and middle-income countries.

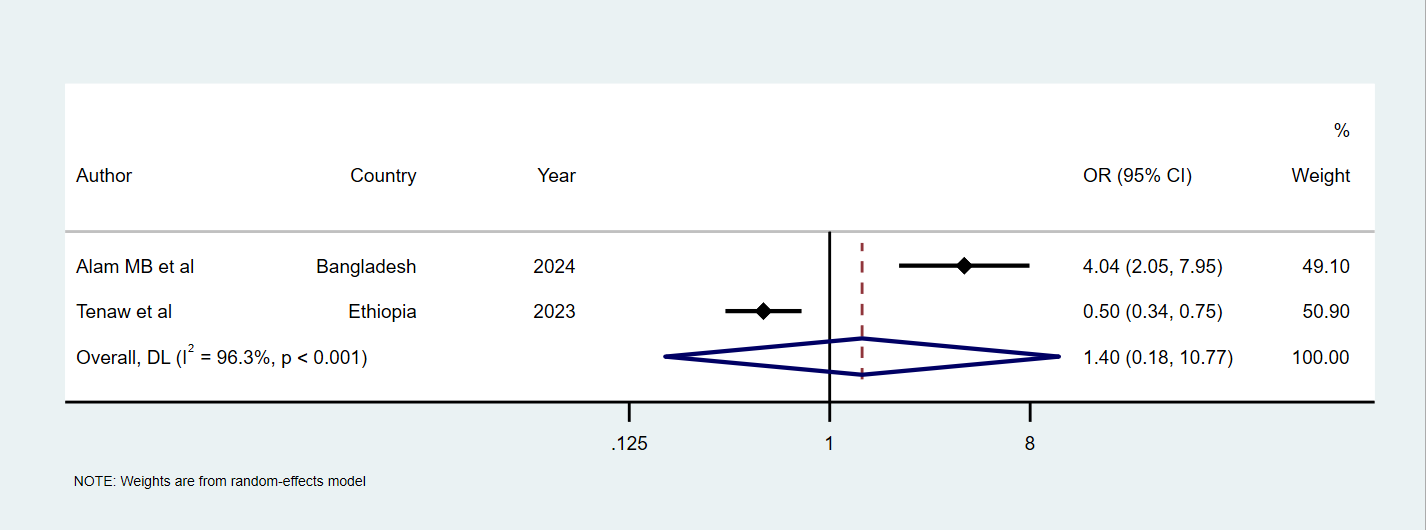

**Supplementary figure 12:** Summary estimations of proximity to healthcare facilities on modern contraception uptake among women with disabilities in low- and middle-income countries.
